# Understanding the Epidemiology of Malaria in Zanzibar through Molecular and Serological Analysis of Samples collected during Reactive Case Detection

**DOI:** 10.1101/2025.05.20.25328026

**Authors:** Varun Goel, Wahida Hassan, Caroline Murphy, Barbara B. Choloi, Mohamed Ali, Bakari Mohamed, Abdallah Zacharia, Msolo C Dominick, Kyaw Thwai, Safia Mohammed, Shija J. Shija, Jeffrey A. Bailey, Anders Björkman, Billy E. Ngasala, Eric Rogier, Jonathan J. Juliano, Jessica T. Lin

**Author notes:** Co-first authors (Varun Goel, Wahida Hassan). Co-senior authors (Jonathan J. Juliano, Jessica T. Lin).

## Abstract

**Background:** The Zanzibar archipelago has been a pre-elimination region for malaria thanks to rigorous control interventions, but recent surges in malaria cases have been observed. The contribution of non-falciparum species to the current malaria situation is unknown.

**Methods:** This study investigates the seroepidemiology of falciparum and non-falciparum malaria species in Zanzibar. Leveraging plasma extracted from dried blood spots (DBS) collected during reactive case detection (RCD) activities across Unguja island from May 2022 to May 2023, we measured immunoglobulin G (IgG) responses to *Plasmodium* MSP1-19kD antigens using a multiplex bead-based immunoassay. Additionally, active infections were detected using species-specific real-time PCR.

**Results:** Out of 1,618 participants surveyed in 35 RCDs, 35.3% had exposure to any malaria species, with *P. falciparum* being the most common (29.8%). Seroprevalences for non-falciparum species were lower: *P. ovale* (5.8%), *P. malariae* (5.9%), and *P. vivax* (5.9%). Active infections were detected in 6.0% of participants, predominantly *P. falciparum* (4.6%). Travel to mainland Tanzania was a dominant risk factor for seropositivity for all four malaria species. Other factors associated with *Pf* seropositivity (high-risk occupations, female status) were not associated with seropositivity for non-falciparum species. The geographic distribution of non-falciparum exposure differed compared to falciparum, with relatively higher seroprevalences in rural districts, especially Kazkazini A in northern Unguja.

**Discussion:** This study suggests a significant contribution of non-falciparum species to the local epidemiology in Zanzibar. Current control and elimination efforts, focused on *P. falciparum*, may not adequately address exposure to non-falciparum species.

## BACKGROUND

The Zanzibar archipelago has been a pre-elimination region for over a decade due to the rigorous implementation of key malaria control interventions including test and treat with artemisinin combination therapies (ACTs), use of reactive case detection (RCD), use of long lasting insecticide treated bed nets (LLINs) and vector control.[1] These interventions have driven malaria prevalence to <1% outside of outbreaks that occur, which increase the number of cases [1]. However, there have been recent surges in malaria cases in the archipelago. In 2020, the annual parasite incidence (API) for Unguja, the main island, reached 11.5 with 12,695 cases being reported [2]. Since then several outbreaks have occurred. Given that *Plasmodium falciparum* is responsible for the vast majority of morbidity and mortality associated with malaria, the majority of resources have gone into its control. However, in nearby mainland Tanzania, as *P. falciparum* has declined, transmission of non-falciparum malaria, in particular *P. ovale* and *P. malariae*, has persisted or even increased, potentially due to differences in how they respond to falciparum malaria control interventions [3]. In certain regions of mainland Tanzania, the prevalence of *P. ovale* can be similar to *P. falciparum* [4]. Given the high levels of connectivity and intense control decreasing falciparum malaria prevalence on the Zanzibar archipelago, we pursued a dedicated investigation of the epidemiology of non-falciparum malaria on the main island of Unguja.

Active infection by malaria can be detected by a number of methods including microscopy, rapid diagnostic tests and nucleic acid testing; the most specific and sensitive of these methods being PCR [5–7]. This is particularly true for non-falciparum malaria, which relies on detection through the lactate dehydrogenase (LDH) band of RDTs which has overall lower sensitivity than histidine rich protein 2 (HRP2) which is specific for falciparum malaria [8]. However, in low transmission settings, active infections may be so infrequent that PCR methods have limited utility. Serological assays can be used to assess prior exposure to malaria in a species specific manner, thus giving a longer term picture of malaria transmission and a better sense of exposure in low transmission settings [9]. To date, there have been sporadic reports of active infection with *P. ovale* [10–12], *P. malariae* [10,12,13], and *P. vivax* [12,13] in Zanzibar. However, there have been no serological surveys of the various malaria species on the archipelago.

Asymptomatic infections that persist and are not treated provide a potential reservoir for malaria transmission[14]. RCDs, typically using malaria rapid diagnostic tests to detect asymptomatic *P. falciparum* cases in the vicinity of the index case identified in clinic, is often employed to find asymptomatic infections, but misses individuals with low density infections. We leveraged dried blood spots (DBS) collected as part of RCD activities with the Zanzibar Malaria Elimination Program (ZAMEP) RCD activities on the island of Unguja, to investigate previous exposure to *P. falciparum*, *P. vivax*, *P. ovale* and *P. vivax* and compare it to rates of active infection detected by real time PCR in asymptomatic individuals. Risk factors for exposure to all four malaria species were further investigated.

## METHODS

### Study design and data collection

We conducted 35 reactive case detection (RCD) visits among the 50 to 100 nearest individuals to an index case across Unguja island in Zanzibar between May 2022 and January 2023, spanning two rounds spaced one month apart. Participants provided DBS, along with travel histories and detailed demographic and household information. The same individuals were not necessarily sampled at each visit and individuals who were sampled twice were only included at the first sampling for analysis. Informed consent/assent was given by all participants or guardians as appropriate. The study was approved by the institutional review boards (IRB) in Zanzibar (ZAHREC/01/EXT/July/2023/03), at MUHAS (MUHAS-REC-02-2024-1168), and at UNC (21-1966). Written informed consent, parental consent and/or assent were provided by all participants.

### Sample processing

Three 6 mm punches were placed in a well of a 96-well deep well plate for DNA extraction, which was performed using a Chelex-Tween extraction protocol [15]. A single 6mm punch was placed in a separate deep well plate for plasma extraction for serology, performed as previously described [16].

### Immunoglobulin G assay analysis and threshold selection

We used a multiplex bead based immunoassay to detect Immunoglobulin G (IgG) against Plasmodium MSP1-19kD antigens to measure exposure to *P. falciparum*, *P*. *malariae*, *P*. *ovale,* and *P. vivax* as previously described [16]. Thresholds of mean fluorescent intensity (MFI) at which an individual is considered seropositive for a particular malaria species were selected using the two-component finite-mixture model (FMM) method [9,16,17]. The FMM method has been applied in previous studies in various settings including Tanzania to quantify malaria exposure in other settings. The FMM method utilizes maximum likelihood estimation approaches to determine two unweighted subpopulations (components) through calculation of their means and variance, yielding two separate distributions for putative seropositive and seronegative individuals for each IgG target. Specifically, the threshold for the analysis was chosen by calculating the *exp(lognormal Mean + 3*Standard Deviations)* of the MSP1-19kD9kD antigen values, similar to the analysis by Rogier et al. [9].

### Real time PCR detection of malaria species

*P. falciparum* parasitemia was determined using a real-time PCR targeting *varATS* as previously described [18,19]. For *P. ovale*, *P. malariae* and *P. vivax* detection, we conducted separate real-time PCR (qPCR) assays for each species, targeting the 18S ribosomal RNA (18S rRNA) gene, enabling both species detection and a semi-quantitative assessment of parasitemia using a standard curve of diluted plasmids for parasitemia estimation (MRA-178, 179 and 180, BEI Resources, Manassas, VA) [15,20]. A genome equivalent was based on a conversion factor of 6 plasmid copies per genome. All plates included negative controls.

### Statistical Analysis

We identified associations between potential geospatial, demographic, and behavioral factors, and previous exposure to falciparum and non-falciparum malaria, by fitting robust poisson regression models using Generalized Estimating Equations (GEEs) to each malaria species, adjusting for individual age and clustering of individuals within the same households. GEE models and FMM estimations were implemented using ‘geepack’ [21], and ‘mixR’ package in R version 4.2.2 [22]. We also conducted sensitivity analyses to assess whether our effect estimates and results are affected by the choice of thresholds based on the FMM method, and whether there may be potential cross-reactivity in IgG responses among the four species-specific *Plasmodium* antigens. Detailed description of the sensitivity analysis and results are provided in the supplement.

## RESULTS

### Study Population

Out of the 1,793 samples collected in the context of two consecutive rounds of reactive case detection (RCD) in the index household and surrounding households, four (0.2%) were excluded due to missing MSP1-19kD IgG data, and 171 (9.5%) were removed as they were associated with participants who had previously participated in the first RCD round. In total, samples from 1,618 participants who participated in 35 RCD rounds from May 2022 to January 2023 were included in the final seroepidemiology analytical dataset. The PCR cohort represented a subset of those included in the seroepidemiology analysis, with 928/1618 (57%) participants sampled from May 2022 to September 2022 also tested for both active falciparum and non-falciparum infections using qPCR.

Overall there were no noticeable differences between the populations included in the sero-epidemiology and qPCR cohorts, although the proportion of qPCR participants was higher in urban settlements as compared to the sero-epidemiology cohort (**Table 1**). For the full seroepidemiology cohort, the median (interquartile range [IQR]) age was 15 (8-29) years, with more female (59%) participants. Most participants lived in urban settlements (72%) in Magharibi B, Magharibi A, and Mjini. Participants mainly consisted of students (44%), followed by housewives (14%), children (13%), and those engaging in trade or business (11%). Among participants that reported their travel history, only 2.7% reported traveling outside of Zanzibar to mainland Tanzania in the last 28 days. Additionally, 30% of households reported having no bednets and a larger majority of participants (59%) reported not sleeping under a bednet the previous night. Most participants (88%) reported living in houses made of brick, cement or both, and almost all participants (98%) lived under a sheet metal, tin, or tiled roof.

**Table 1:** Baseline characteristics of participants in the seroprevalence and qPCR cohorts.

| Variable | Seroprevalence |  | qPCR |  |
| --- | --- | --- | --- | --- |
|  | N | (%) | N | (%) |
| **Age** | 1618 |  | 927 |  |
| <8 | 344 | (21.3) | 203 | (21.9) |
| 8 to 15 | 469 | (29.0) | 263 | (28.4) |
| 16 to 30 | 436 | (26.9) | 246 | (26.5) |
| >30 | 369 | (22.8) | 215 | (23.2) |
| **Sex** | 1618 |  |  |  |
| Female | 957 | (59.1) | 533 | (57.5) |
| Male | 661 | (40.9) | 394 | (42.5) |
| **Settlement Type** |  |  |  |  |
| Urban | 1171 | (72.4) | 802 | (86.5) |
| Rural | 447 | (27.6) | 125 | (13.5) |
| **District** | 1618 |  |  |  |
| Magharibi B (urban and rural) | 495 | (30.6) | 311 | (33.5) |
| Magharibi A (urban) | 380 | (23.5) | 265 | (28.6) |
| Mjini (urban) | 380 | (23.5) | 226 | (24.4) |
| Kati (rural) | 180 | (11.1) | 42 | (4.5) |
| Kusini (rural) | 99 | (6.1) | 48 | (5.2) |
| Kaskazini A (rural) | 84 | (5.2) | 35 | (3.8) |
| **Occupation** | 1608 |  |  |  |
| Student | 704 | (43.8) | 421 | (45.8) |
| Housewife | 225 | (14.0) | 119 | (12.9) |
| Child | 216 | (13.4) | 113 | (12.3) |
| Trader/Business | 179 | (11.1) | 99 | (10.8) |
| Farming | 73 | (4.5) | 35 | (3.8) |
| Fishing | 66 | (4.1) | 45 | (4.9) |
| Teacher | 23 | (1.4) | 13 | (1.4) |
| Retired or Unemployed | 19 | (1.2) | 12 | (1.3) |
| Public servant/NGO | 16 | (1.0) | 8 | (0.9) |
| Food service | 11 | (0.7) | 6 | (0.7) |
| Watchman | 10 | (0.6) | 2 | (0.2) |
| Construction | 8 | (0.5) | 0 | (0.0) |
| Tourism | 6 | (0.4) | 2 | (0.2) |
| Other | 52 | (3.2) | 37 | (3.9) |
| **Number of bed nets in home** | 1618 |  |  |  |
| 0 | 487 | (30.1) | 316 | (34.1) |
| 1 to 2 | 469 | (29.0) | 281 | (30.3) |
| 3+ | 662 | (40.9) | 330 | (35.6) |
| **Age of bed nets** | 1107 |  | 612 |  |
| <1 year | 639 | (57.7) | 372 | (60.8) |
| 1 to 2 years | 426 | (38.5) | 214 | (35.0) |
| 3+ years | 42 | (3.8) | 26 | (4.2) |
| **Slept under bed net last night?** | 1588 |  | 914 |  |
| Yes | 651 | (41.0) | 370 | (40.5) |
| No | 937 | (59.0) | 544 | (59.5) |
| **Indoor residual spraying** |  |  | 911 |  |
| Yes | 369 | (23.7) | 259 | (28.4) |
| No | 1190 | (76.3) | 652 | (71.6) |
| **Last indoor residual spraying** | 353 |  | 249 |  |
| <6 months | 40 | (11.3) | 38 | (15.3) |
| 6 to <12 months | 280 | (79.3) | 190 | (76.3) |
| >12 months | 33 | (9.3) | 21 | (8.4) |
| **House Material** | 1577 |  | 916 |  |
| Bricks & Cement | 767 | (53.1) | 501 | (54.7) |
| Cement | 483 | (33.4) | 253 | (27.6) |
| Bricks | 136 | (9.4) | 50 | (5.5) |
| Stone with mud | 58 | (4.0) | 41 | (4.4) |
| Mud | 13 | (0.9) | 10 | (1.1) |
| Bamboo with mud | 11 | (0.8) | 11 | (1.2) |
| Other (combination) | 109 | (7.5) | 50 | (5.5) |
| **Roof** | 1479 |  | 864 |  |
| Sheet metal/tin/tiles | 1455 | (98.4) | 848 | (98.2) |
| Thatched/leaves/bamboo | 24 | (1.6) | 16 | (1.8) |
| **Water Source within 50 meters of house?** | 1512 |  | 903 |  |
| Yes | 189 | (12.5) | 113 | (12.5) |
| No | 1251 | (82.7) | 748 | (82.8) |
| Don't know | 72 | (4.8) | 42 | (4.7) |
| **Travel** | 1540 |  | 919 |  |
| None | 1498 | (97.3) | 911 | (99.1) |
| Mainland Tanzania | 42 | (2.7) | 8 | (0.9) |

### Seroprevalence Estimates

Out of the 1,618 Zanzibar residents tested for anti-MSP1-19kD IgG antibody responses, 35% of participants (n = 571/1617) had evidence for exposure to any malaria species. The most common exposure was *P. falciparum* with 30% of participants (n = 482/1617) being seropositive. Seroprevalence for each non-falciparum malaria species was much lower; *P. ovale* was 5.8% (n=94/1611), while *P. malariae* and *P. vivax* seroprevalence were 5.9% (n = 96/1617 and n = 95/1617) each (**Figure 1**). Among the participants with seroprevalence results available for all four malaria species (n =1611), exposure to multiple malaria species was common with 133 (23% out of 570) having exposure to two or more species, and 15 (2.8%) with evidence of exposure to all four species. Overall, 36% (203/570) of the malaria-exposed group demonstrated exposure to non-falciparum species; nearly half of these (44%, 89/203) showed exposure to non-falciparum species in the absence of exposure to falciparum.

**Figure 1:**
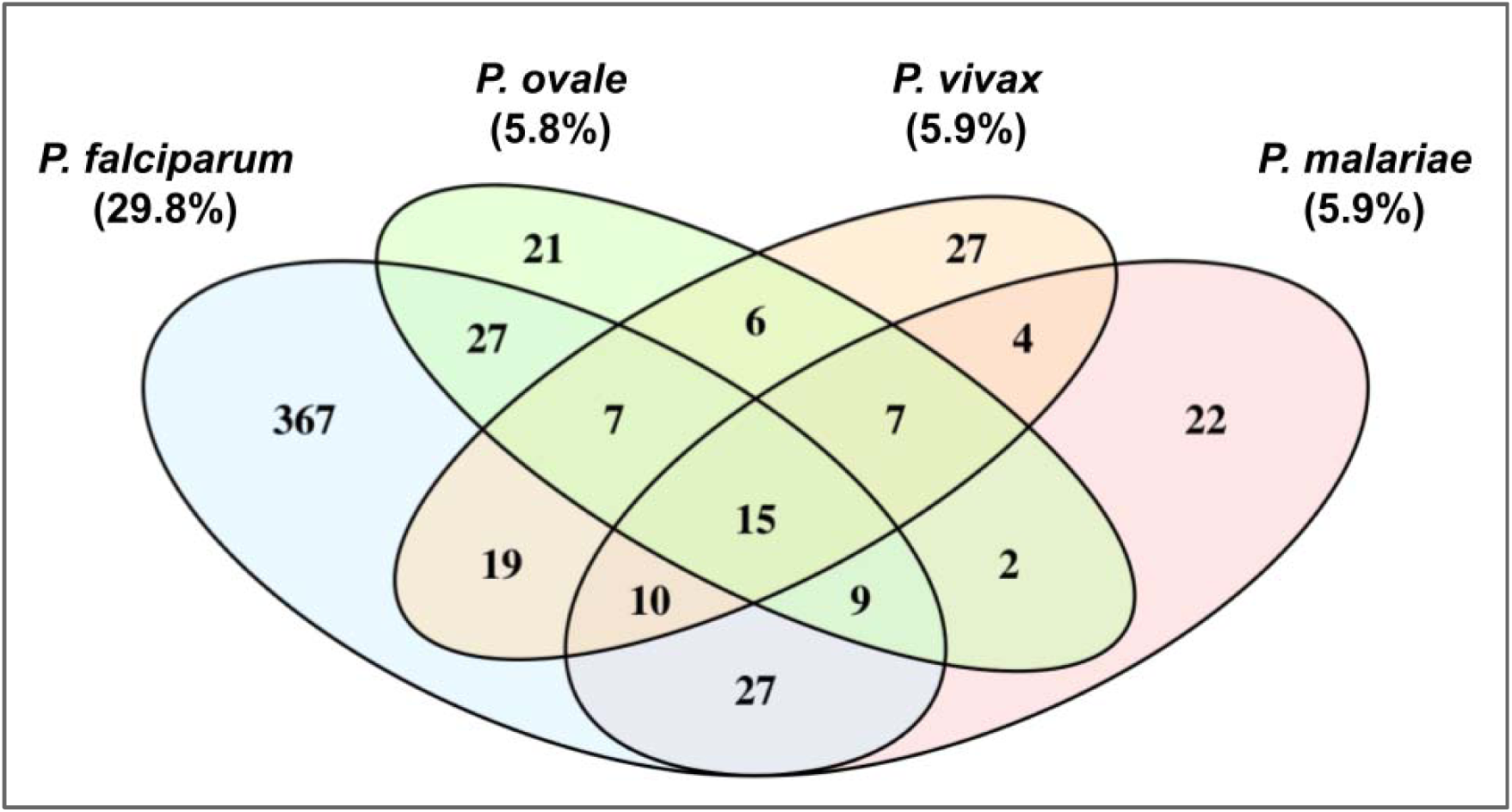
Overlap of seroprevalence across all four malaria species. In total, IgG vs MSP1-19kD responses were measured across 1,618 Zanzibar residents participants in reactive case detection after a malaria case was identified in proximity (in their or a neighboring household). Note: Percentages for *P. ovale* were calculated for 1,611 individuals with successful seroprevalence results. Percentages for other species were calculated for 1,617 individuals. 1,611 participants had seroprevalence results available for all 4 species.

We conducted a sensitivity analysis by adopting a very stringent threshold at 5 SD instead of 3 SD, obtaining seroprevalence estimates for *P. falciparum*, *P. ovale*, *P. malariae*, and *P. vivax* seroprevalence of 24%, 2.4%, 3.0%, and 2.0%, respectively (**Supplementary Table 2** and **3**). In this analysis, 23% (100/429) of the malaria-exposed group demonstrated exposure to non-falciparum species; nearly half of these (42%, 42/100) showed exposure to non-falciparum species in the absence of exposure to falciparum. To assess for potential cross-reactivity responses across antigens, we also examined correlation of MFI responses across all four MSP1-19kD orthologous antigens **(Supplementary** Figure 2), and the distributions in MFI responses for individuals with mono-species vs mixed-species exposure (**Supplementary** Figure 3). Although persons seropositive against multiple species had slightly higher MFIs to *Po* and *Pm* MSP1-19kD antigens than those seropositive against only one species-specific antigen, those differences were not significant.

We also used our cross-sectional data to estimate the cumulative proportion of exposure over the lifespan to understand when exposure is usually acquired (**Figure 2**). Overall, approximately 30% of the population was exposed to *P. falciparum*, with a steady increase in the proportion of persons seropositive over childhood years. By age 28 years, approximately 15% (half the total seroprevalence) of individuals showed exposure. For non-falciparum infection, the rate of acquisition was similar between species and perhaps slightly faster than for *P. falciparum*, with half of those exposed (3%) by the age of 20 years.

**Figure 2.**
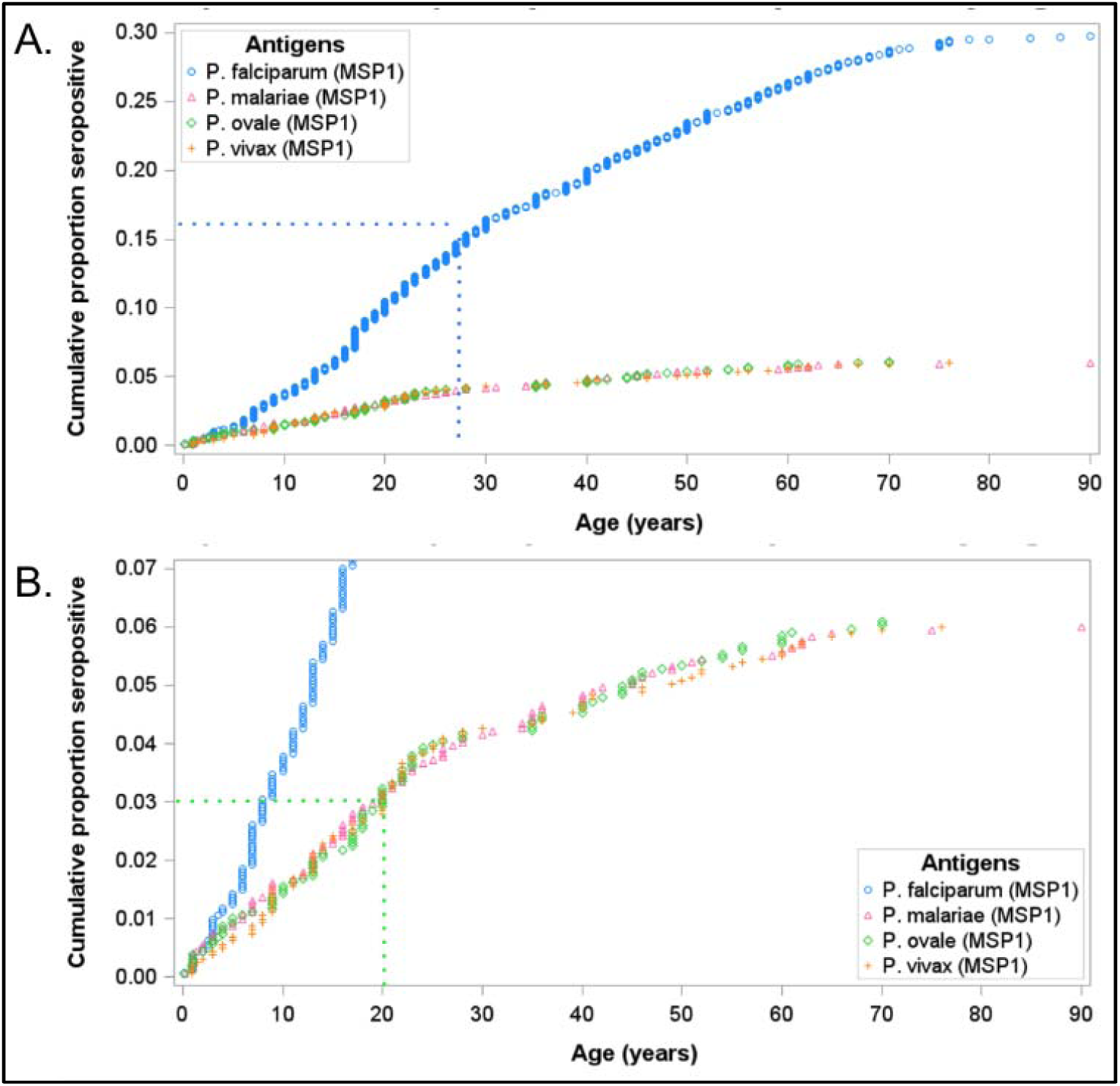
Cumulative exposure to malaria. Panel A shows the overall cumulative exposure to all 4 malaria species tested. Panel B is a limited view at lower proportions to assess non-falciparum cumulative exposure.

### PCR testing and active infections

As expected, compared to malaria seroprevalence estimates, rates of active infection were much lower. Overall, 56/928 (6.0%) were infected with any malaria species. 43 (4.6%) tested positive for *P. falciparum* parasitemia, with 77% of those participants (n = 33) having very low parasite density of <1 parasite/μL (**Supplementary** Figure 2). Similarly, few active non-falciparum infections were detected; *P*. *malariae* parasitemia was detected in 11 (1.2%), *P*. *ovale* in 4 (0.4%), and *P. vivax* in only 2 (0.2%) participants, respectively. Among these cases, only 2 reported recent travel to the mainland, to the Simayu region, northwest Lake Zone (*P. malariae* case), and to Dar es Salaam (*P. ovale* case). Both *P. vivax* parasitemias were detected in the same household, in a housewife and a 4 month-old infant, both Zanzibari residents without recent travel.

### Factors associated with seropositivity

Given the low rate of active infection by qPCR, especially for non-falciparum species, we examined potential factors associated with prior malaria exposure. In this exploratory analysis, demographic factors associated with seropositivity showed differing trends for falciparum vs. non-falciparum malaria (**Figure 3** and **Supplementary Table 1**). For *P. falciparum*, seroprevalence among persons greater than 15 years of age was 5.1 (95% CI: 3.0 – 8.7) times compared to children under five years of age. In contrast, for *P. ovale*, children between 5 to 15 years of age had half the seroprevalence (PR = 0.49, 95% CI: 0.25 – 0.97) of *P. ovale* compared to children under five years of age. Females, compared to males, had higher seroprevalence ratios for *P. falciparum* (PR = 1.20, 95% CI: 1.05 – 1.38) and *P. ovale* (PR = 1.71, 95% CI: 1.08 – 2.71), while controlling for age and clustering of individuals within households. Compared to students, occupations such as farming, trading and fishing, or being a housewife were associated with higher seroprevalence for *P. falciparum*, but not for any non *P. falciparum* species. However, associations among these demographic factors and non-falciparum malaria were not significant after assuming a stringent 5 SD threshold for seropositivity (**Supplementary** Figure 5).

**Figure 3.**
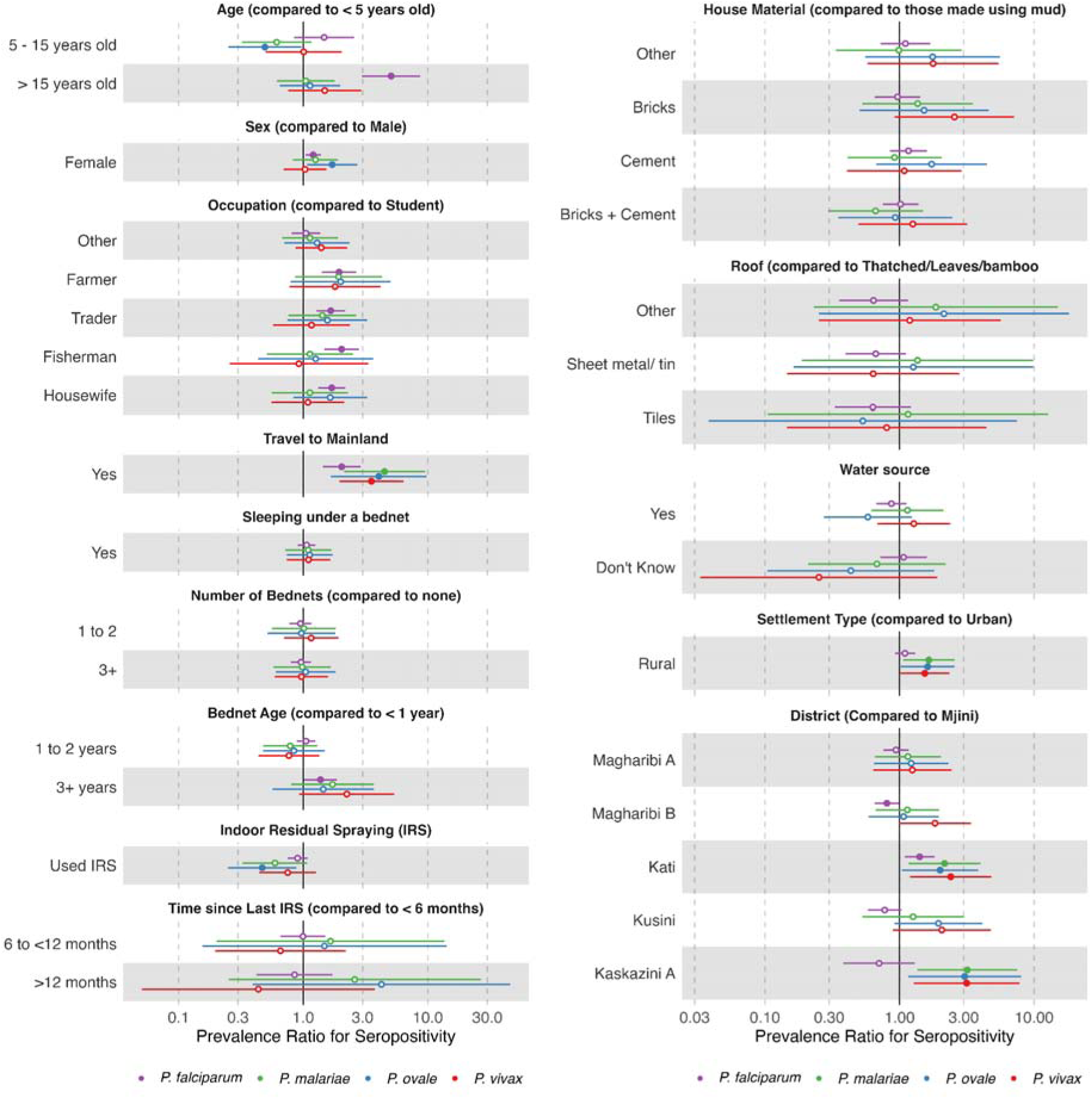
Factors associated with seropositivity for all four species. Seroprevalence ratios were estimated by fitting robust poisson regression models using Generalized Estimating Equations (GEEs) to each malaria species, adjusting for individual age and clustering of individuals within the same households.

Overall, travel to mainland Tanzania in the last 28 days was the most strongly associated with high seropositivity for all four species. Travel to mainland Tanzania was associated with twice the seroprevalence for *P. falciparum* (PR = 2.0; 95% CI: 1.4 – 2.9), and upwards of three times the seroprevalence for all non-falciparum malaria species (**Figure 3**). While sleeping under a bednet the previous night or increasing number of bednets in a household was not associated with seroprevalence for both falciparum and non-falciparum species, increasing bednet age was associated with higher *P. falciparum* exposure (PR = 1.37 (95% CI: 1.01 - 1.87) in households that had bednets older than three or more years compared to participants in households with less than a year-old bednets). Indoor residual spraying (IRS) was associated with a protective effect only for *P. ovale* seroprevalence (PR = 0.47, 95% CI: 0.25 – 0.88). Household construction materials, roof type, and water source were not associated with seropositivity for any species. In our sensitivity analysis assuming a 5SD threshold, these associations persisted, with the exception of the association between IRS and *P. ovale* seroprevalence **(Supplementary** Figure 5).

### Geographic distribution of malaria exposure

We also observed some geographic differences in seroprevalence across Unguja for falciparum and non-falciparum malaria. Rural areas were associated with higher seroprevalence for all three non-falciparum species, but not with *P. falciparum* (**Figure 3**). Additionally, districts with the highest *P. falciparum* seroprevalence were not the same as districts with the highest seroprevalence for each non-falciparum species (**Figure 4**). Highest *P. falciparum* seroprevalence was found in the central district of Kati followed by the western district of Mjini, while higher non-falciparum seroprevalence (*P. malariae*, *P. ovale*, and *P. vivax*) was concentrated in the Kaskazini A district in northern Unguja. Models controlling for age and household clustering also show that when compared to an urban district such as Mjini, non-falciparum seroprevalence ratios were significantly higher for rural districts such as Kati and Kaskazini A, but not other urban districts. Even if we assume a threshold of 5 SD for defining seroprevalence, the positive associations between non-falciparum seroprevalence and rurality existed, especially for *P. malariae*, although the confidence intervals crossed the null for other non-falciparum species **(Supplementary** Figure 5). However, we also note that overall *P. falciparum* seroprevalence within the districts of Unguja is much higher (ranging from 25% to 35%), compared to non-falciparum species (ranging from 6% to 14%).

**Figure 4:**
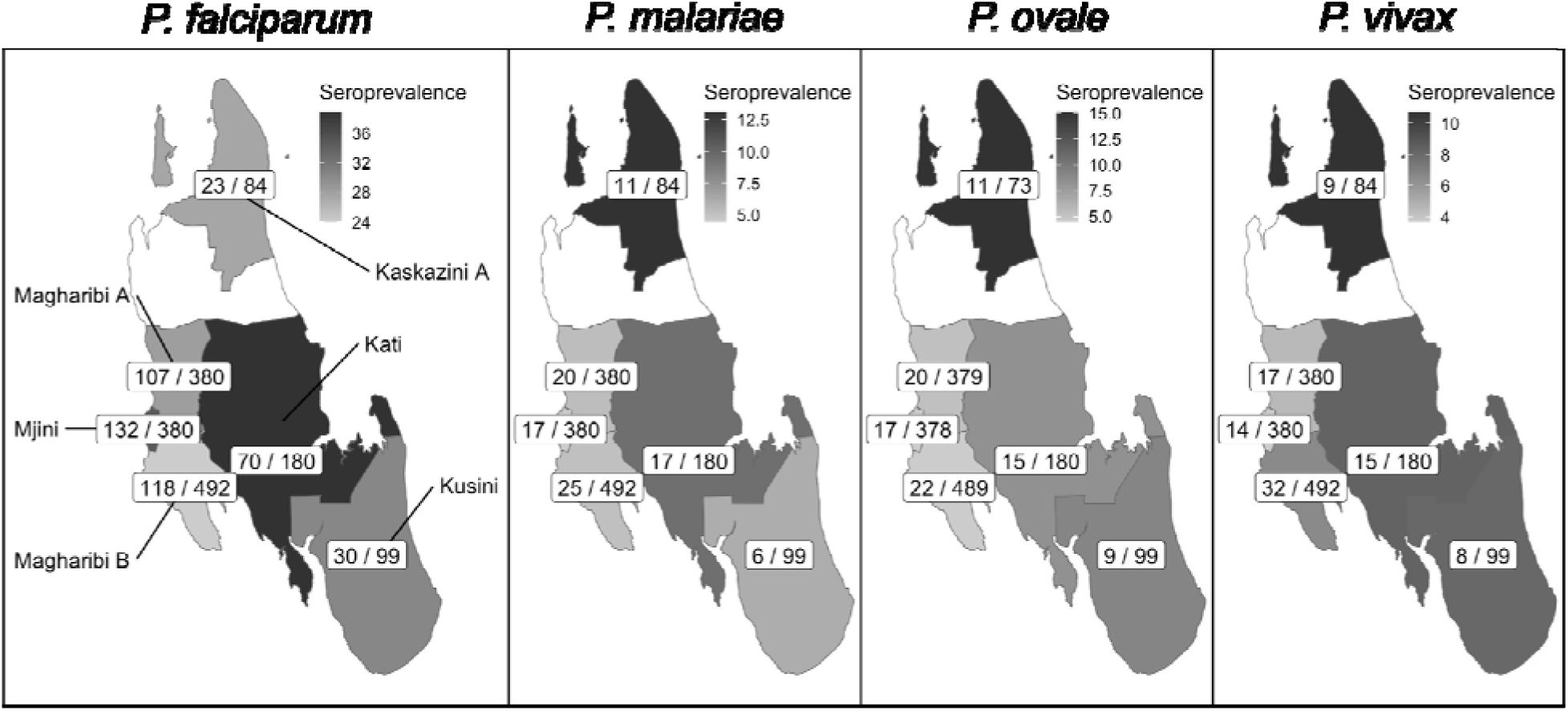
Map of geographic distribution of seroprevalence across all four species. Each district is shaded based on the species specific sero-prevalence. Within each district are the number of seropositive samples (# positive/# tested)

## DISCUSSION

We conducted a comprehensive seroepidemiology study of the four human malaria species in Zanzibar and compared exposure to active infection detected by qPCR in asymptomatic individuals. After nearly two decades of maintaining pre-elimination status, exposure to *P. falciparum* remains common in the population (30%), perhaps attesting to both the longevity of IgG responses as well as ongoing malaria exposure, whether on Zanzibar or elsewhere. The longevity of anti-MSP1-19kD IgG responses varies by age and type of stimulus, but this is known to be a highly-immunogenic malaria antigen with durable IgG levels in plasma for years or decades [23,24]. Cumulative prevalence of exposure increased with age, with about 50% of exposure occurring in the first 3 decades of life. Serological tools to measure (possibly distant) previous malaria exposure are especially important in a low-transmission setting - and emphasized by the rates of active infection in this study with only 4.3% of individuals infected with *P. falciparum*, mostly at very low density, under <1 parasite/μL blood.

Exposure to non-falciparum infection was lower but significant, and similar between the three species tested at approximately 6%. Cumulative exposure for these species reached its midpoint slightly earlier, around 20 years of age. Though active infections were very uncommon, they were more prevalent than previously seen in a large PCR survey conducted six years earlier, in 2016 [25]. Though only 2.7% of the study population reported traveling outside of Zanzibar to mainland Tanzania in the last 28 days, exposure to non-falciparum infection was significantly higher in travelers compared to those not reporting recent travel. We did not collect data on more remote travel, and it is possible that the acute infection that led to an IgG response occurred outside of Zanzibar. *P. malariae*, *P. ovale*, and *P. vivax* all have biological mechanisms that can lead to persistent or chronic latent infection that may support maintenance of detectable IgG levels even if exposure was more remote.

The similarity of seroprevalence among non-falciparum species might raise concerns about cross reactivity of IgG responses to orthologous MSP1-19kD antigens. Several of our analyses support that these signals represent true exposure. In this study, MFI intensity did not seem to vary with co-infection. Cross reactivity between antibodies generated to a falciparum exposure and a non-falciparum antigen would likely present as lower non-falciparum MFI in the co-infections. Also, no specific patterns of correlation were determined. These antigens have been used extensively for sero-epidemiology studies and the risk of cross-reactivity has been studied and deemed to be low [17]. Given this, the reported seroprevalence is likely to be representative of exposure in this population. Applying even more stringent cutoffs for seropositivity still yielded evidence of exposure to each of the non-falciparum species in the study population.

Known risk factors were identified for *P. falciparum* exposure, including being a school aged child. Significantly higher seroprevalence among those greater than 15 years of age may also potentially be explained by the fact that those populations may have been exposed to falciparum before widespread implementation of IRS and long lasting insecticidal nets (LLINs) in 2007-2008 in Zanzibar [1,26]. Interestingly, the risk for *P. ovale* seropositivity was higher in younger children (under 5 years) than older children (5-15 years). This suggests a recent exposure in the population. Younger children may be exposed when traveling with their parents to the mainland, as we have seen that presumptive imported cases may disproportionately affect childrenL<L5Lyears and housewives compared with nonLtravel–related cases (unpublished data).

The relative geographic distribution of exposure to the different malaria species was not the same. *P. falciparum* exposure was highest in the Kati district in the central-East portion of the island. Kati has had relatively high annual parasite incidence (API) in the years leading into this study, with an API of 18.3 in 2020 and 11.6 in 2021 [2]. In 2022, the API was lower (5.3) which may in part be reflected in the infrequently detected active infections by qPCR during the study (43/928). The seroprevalence of all three non-falciparum species was highest in Kaskazini A, an area with lower transmission with APIs of 7.6, 3.3 and 1.7 in 2020, 2021 and 2022, respectively [2]. This is not surprising, as studies of active infection by non-falciparum malaria in Tanzania have shown elevated risks in regions of lower transmission [3,4,15].

Limitations to this study include its cross-sectional nature and possible bias towards increased malaria exposure compared to the general population, based on its reactive case detection study design. This sampling strategy targets asymptomatic persons who were at home during the visit by the malaria surveillance officer, and therefore does not address the epidemiology of symptomatic malaria and likely under-samples populations engaged in occupational activities outside the home. Our study was not large enough to perform fully adjusted multivariate analyses for factors associated with seropositivity. While the longevity of anti-MSP1-19kD IgG responses has been studied for *P. falciparum*, much less is known about the longevity of response to the orthologous antigen in non-falciparum species. Additionally, despite the increased sensitivity of PCR for detecting non-falciparum species, the assays used likely still miss the lowest density parasitemias that can be found in these species. Finally, given the possibility of persistent, latent, and relapsed infection, we are unable to determine whether the few active parasitemias of non-falciparum species were locally acquired.

Despite relatively low transmission, exposure to malaria, in particular falciparum malaria, remains high in Zanzibar with over a third (36%) of the population being seroreactive to any malaria species. Immediately prior to these collections, there was a major surge of malaria in 2021, potentially highlighted by the high seroprevalence but low level of active infection during 2022-2023. Sero-reactivity to non-falciparum infections was also prevalent, including to *P. vivax* which has historically been low in Africa [27]. We provide new seroepidemiological data to suggest these species are important to the local epidemiology in Zanzibar. Further research is needed to better understand the context of exposure to non-falciparum malaria species, whether *P. ovale*/*vivax* seropositive individuals are at risk for relapse, and whether local conditions are conducive to local transmission of non-falciparum species. Beyond targeting travelers, it is unclear whether current elimination efforts focused on *P. falciparum* will adequately address exposure to non-falciparum species.

## Supporting information

Link to Supplement

## Acknowledgements

We thank the study participants who donated their time and the health facility staff who collected samples for the study. We thank the ZAMEP district malaria surveillance officers who were instrumental to the successful implementation of the study. The following reagents were obtained through BEI Resources, NIAID, NIH: Diagnostic Plasmid Containing the Small Subunit Ribosomal RNA Gene (*18S*) from *Plasmodium vivax*, MRA-178; *Plasmodium malariae*, MRA-179; and *Plasmodium ovale*, MRA-180, contributed by Peter A. Zimmerman.

## Author Contributions

Conception and design: JJJ, JTL, ER; Data acquisition, analysis and interpretation: VG, WH, CM, BC, MA, BM, AZ, MD, KT, SM, SS, JAB, AB, BEN, ER, JJJ, JTL; Drafting manuscript: VG, JJJ, JTL; Revision and final approval: All authors; Accountability: VG, JJJ, JTL

## Data Availability

Metadata are available upon request to the corresponding author.

