## Supplementary material for "Understanding the Epidemiology of Malaria in Zanzibar through Molecular and Serological Analysis of Samples collected during Reactive Case Detection": Link to Supplement

Supplementary Materials………………………………………………………………..	2

Supplementary Table 1………………………………………………………………….	3

**SUPPLEMENTAL MATERIALS**

**Concordance between qPCR and Seroprevalence Estimates**

Among participants that tested positive for active falciparum infections (positive PCR), 56% (24/43) also tested positive for previous *P. falciparum* exposure. Among the 11 participants with active *P*. *malariae* infections, two participants were also considered IgG positive to previous *P*. *malariae* exposure. None of the other participants with active *P.* *ovale spp.* or *P. vivax* infections tested positive for previous *P*. *ovale* or *P*. *vivax* exposure.

**Sensitivity Analysis**

We also conducted a sensitivity analysis to examine the impact of various cutoff values on estimates of seroprevalence for all four malaria species (**Supplementary Table 2** and **Table 3**). Additionally, we also compared the IgG assay signal with each species' MSP1-19kD antigen compared to all others to examine potential for cross reactivity (**Supplementary Figure 2**). We further assess potential for cross-reactivity by examining distribution of MSP1-19kD antigen values for each species by comparing seropositive individuals with exposure to one species and those with exposure to multiple species (**Supplementary Figure 3**) [[1]](https://paperpile.com/c/dYNeb1/2nLH). These distributions were also compared for the most stringent cutoff values for seropositivity (**Supplementary Figure 4**). Lastly, we also examined whether associations between seroexposure and relevant demographic, behavioral or geographic factors were affected by the choice of cutoffs for seropositivity (**Supplementary Figure 5**).

**Supplementary Table 1: Distribution of seroprevalence across baseline characteristics and four species**

| **Variable** | ***P. falciparum*** | | | ***P. malariae*** | | | ***P. ovale*** | | | ***P. vivax*** | | |
| --- | --- | --- | --- | --- | --- | --- | --- | --- | --- | --- | --- | --- |
|  | n/N | (%) | *p*-value | n/N | (%) | *p*-value | n/N | (%) | *p*-value | n/N | (%) | *p*-value |
| **Age** |  |  |  |  |  |  |  |  |  |  |  |  |
| <5 years | 19/188 | (10.1) | <0.001 | 13/188 | (6.9) | 0.028 | 14/187 | (7.5) | <0.001 | 9/188 | (4.8) | 0.127 |
| 5 to 15 years | 82/623 | (13.2) |  | 25/623 | (4.0) |  | 20/621 | (3.2) |  | 30/623 | (4.8) |  |
| >15 years | 379/804 | (47.1) |  | 59/804 | (7.3) |  | 64/801 | (8.0) |  | 58/804 | (7.2) |  |
| **Sex** |  |  |  |  |  |  |  |  |  |  |  |  |
| Female | 301/954 | (31.6) | 0.053 | 63/954 | (6.6) | 0.225 | 68/949 | (7.2) | 0.031 | 58/954 | (6.1) | 0.881 |
| Male | 179/661 | (27.1) |  | 34/661 | (5.1) |  | 30/660 | (4.5) |  | 39/661 | (5.9) |  |
| **Settlement Type** |  |  |  |  |  |  |  |  |  |  |  |  |
| Urban | 337/1168 | (28.9) | 0.217 | 60/1168 | (5.1) | 0.018 | 61/1162 | (5.2) | 0.023 | 62/1168 | (5.3) | 0.056 |
| Rural | 143/447 | (32.0) |  | 37/447 | (8.3) |  | 37/447 | (8.3) |  | 35/447 | (7.8) |  |
| **Occupation** |  |  |  |  |  |  |  |  |  |  |  |  |
| Student | 112/702 | (16.0) | <0.001 | 34/702 | (4.8) | 0.417 | 32/702 | (4.6) | 0.114 | 34/702 | (4.8) | 0.104 |
| Housewife | 110/224 | (49.1) |  | 15/224 | (6.7) |  | 19/222 | (8.6) |  | 14/224 | (6.3) |  |
| Child | 22/216 | (10.2) |  | 13/216 | (6.0) |  | 10/214 | (4.7) |  | 10/216 | (4.6) |  |
| Trader/Business | 87/179 | (48.6) |  | 14/179 | (7.8) |  | 13/178 | (7.3) |  | 12/179 | (6.7) |  |
| Farming | 49/73 | (67.1) |  | 8/73 | (11.0) |  | 8/73 | (11.0) |  | 8/73 | (11.0) |  |
| Fishing | 37/66 | (56.1) |  | 4/66 | (6.1) |  | 5/66 | (7.6) |  | 4/66 | (6.1) |  |
| Other | 63/145 | (43.4) |  | 9/145 | (6.2) |  | 11/144 | (7.6) |  | 15/145 | (10.3) |  |
| **No. bed nets in home** |  |  |  |  |  |  |  |  |  |  |  |  |
| 0 | 149/487 | (30.6) | 0.838 | 29/487 | (6.0) | 0.982 | 30/486 | (6.2) | 0.982 | 29/487 | (6.0) | 0.637 |
| 1 to 2 | 140/469 | (29.9) |  | 29/469 | (6.2) |  | 29/468 | (6.2) |  | 32/469 | (6.8) |  |
| 3+ | 191/659 | (29.0) |  | 39/659 | (5.9) |  | 39/655 | (6.0) |  | 36/659 | (5.5) |  |
| **Age of bed nets** |  |  |  |  |  |  |  |  |  |  |  |  |
| <1 year | 180/636 | (28.3) | 0.028 | 41/636 | (6.4) | 0.162 | 39/632 | (6.2) | 0.479 | 39/636 | (6.1) | 0.029 |
| 1 to 2 years | 130/426 | (30.5) |  | 21/426 | (4.9) |  | 22/425 | (5.2) |  | 19/426 | (4.5) |  |
| 3+ years | 20/42 | (47.6) |  | 5/42 | (11.9) |  | 4/42 | (9.5) |  | 6/42 | (14.3) |  |
| **Indoor residual spraying** |  |  |  |  |  |  |  |  |  |  |  |  |
| Yes | 95/369 | (25.7) | 0.062 | 14/369 | (3.8) | 0.061 | 12/369 | (3.3) | 0.011 | 17/369 | (4.6) | 0.204 |
| No | 366/1187 | (30.8) |  | 76/1187 | (6.4) |  | 81/1181 | (6.9) |  | 76/1187 | (6.4) |  |
| **Roof** |  |  |  |  |  |  |  |  |  |  |  |  |
| Sheet metal/tin/tiles | 432/1452 | (29.8) | 0.411 | 86/1452 | (5.9) | 0.717 | 84/1446 | (5.8) | 0.733 | 86/1452 | (5.9) | 0.621 |
| Thatched/leaves/bamboo | 9/24 | (37.5) |  | 1/24 | (4.2) |  | 1/24 | (4.2) |  | 2/24 | (8.3) |  |
| **Travel** |  |  |  |  |  |  |  |  |  |  |  |  |
| None | 433/1495 | (29.0) | <0.001 | 82/1495 | (5.5) | <0.001 | 82/1489 | (5.5) | <0.001 | 83/1495 | (5.6) | <0.001 |
| Mainland Tanzania | 26/42 | (61.9) |  | 10/42 | (23.8) |  | 9/42 | (21.4) |  | 8/42 | (19.0) |  |

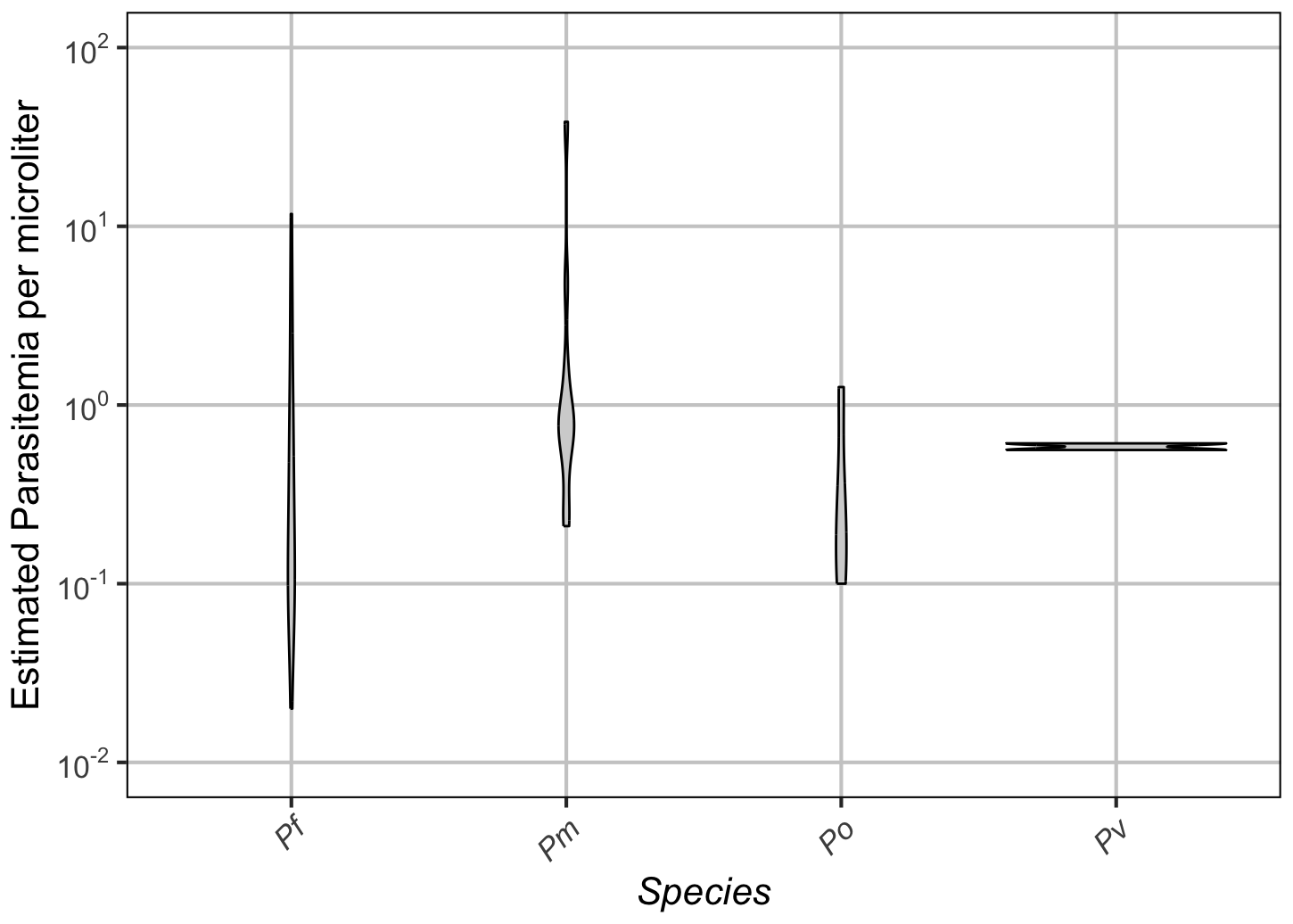

**Supplementary Figure 1. PCR parasitemia determined by real time PCR.** The distribution of estimated parasitemias for PCR positive samples for each species is shown. Pf: *P. falciparum*; Pv: *P. vivax*; Pm: *P. malariae*; Po: *P. ovale spp*. Results are shown on a log_10_ scale.

**Supplementary Table 2: Seropositivity cutoff values based on various standard deviations using two components Finite Mixture Models (FMM).** Cutoffs for the analysis were chosen by calculating the exp(lognormal Mean + 3*Standard Deviations) of the MSP-19 antigen values, similar to the analysis by [[2]](https://paperpile.com/c/dYNeb1/dk3H).

| **Species** | **1 SD** | **2 SD** | **3 SD** | **4 SD** | **5 SD** |
| --- | --- | --- | --- | --- | --- |
| *P. falciparum* | 6.1 | 11.1 | 20.4 | 37.4 | 68.7 |
| *P. ovale spp.* | 15.9 | 30.7 | 59.6 | 115.6 | 224.1 |
| *P. malariae* | 8.4 | 16.6 | 32.8 | 64.9 | 128.4 |
| *P. vivax* | 10.3 | 20.9 | 42.4 | 86.1 | 174.6 |

**Supplementary Table 3: Distribution of seroprevalence (in %) at different seropositivity thresholds for all malaria species.**

| **Species** | **1 SD** | **2 SD** | **3 SD** | **4 SD** | **5 SD** |
| --- | --- | --- | --- | --- | --- |
| *P. falciparum* | 41.4 | 34.0 | 29.8 | 26.5 | 24.0 |
| *P. ovale spp.* | 24.3 | 11.1 | 5.8 | 3.6 | 2.4 |
| *P. malariae* | 22.5 | 11.1 | 5.9 | 4.2 | 3.0 |
| *P. vivax* | 23.6 | 11.8 | 5.9 | 3.5 | 2.0 |

**Supplementary Figure 2. Scatterplots of IgG assay signal with each species' MSP1-19kD antigen compared to all others.** Red line shows linear regression fitting, with inset text
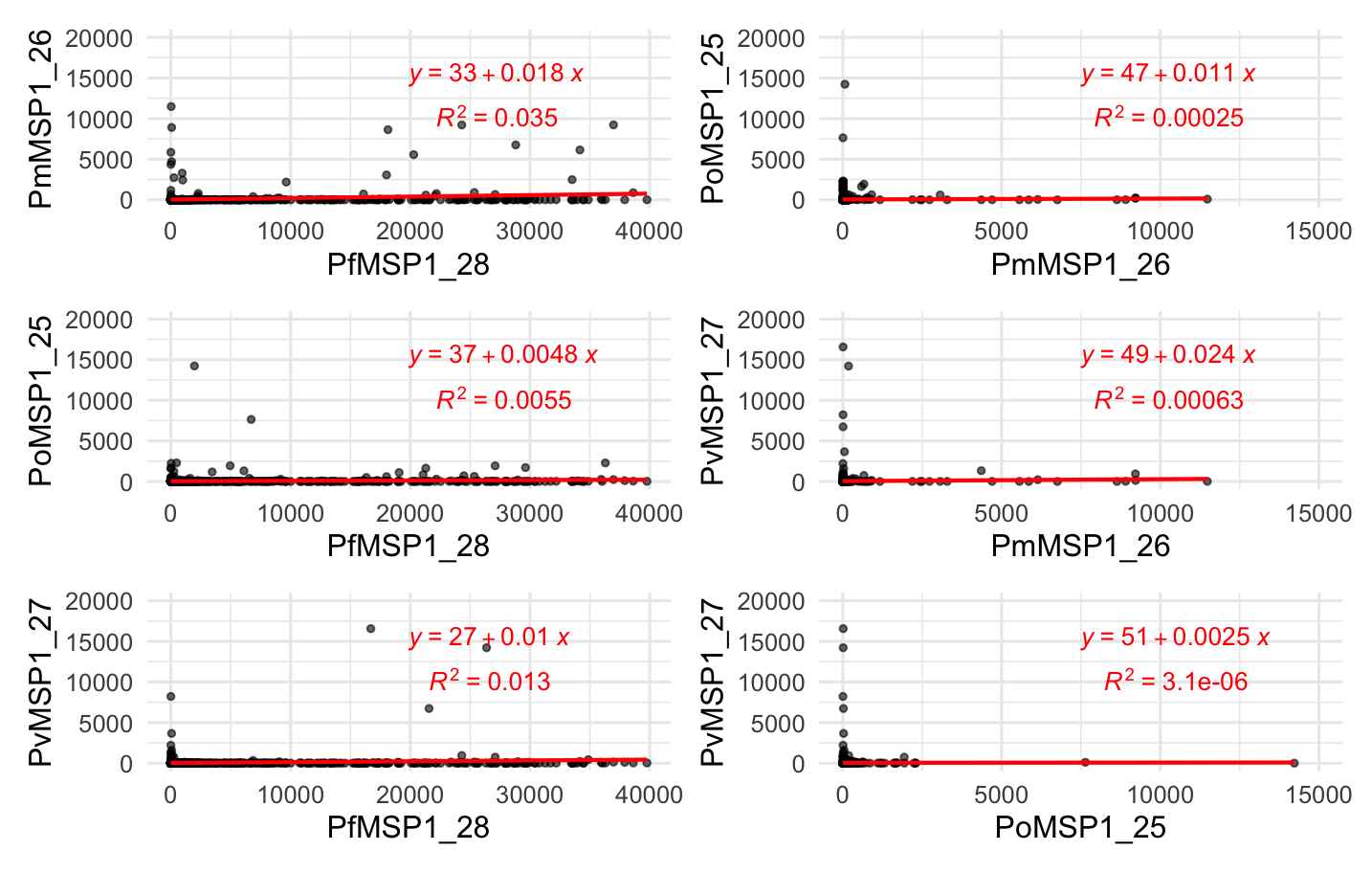

displaying model estimates.

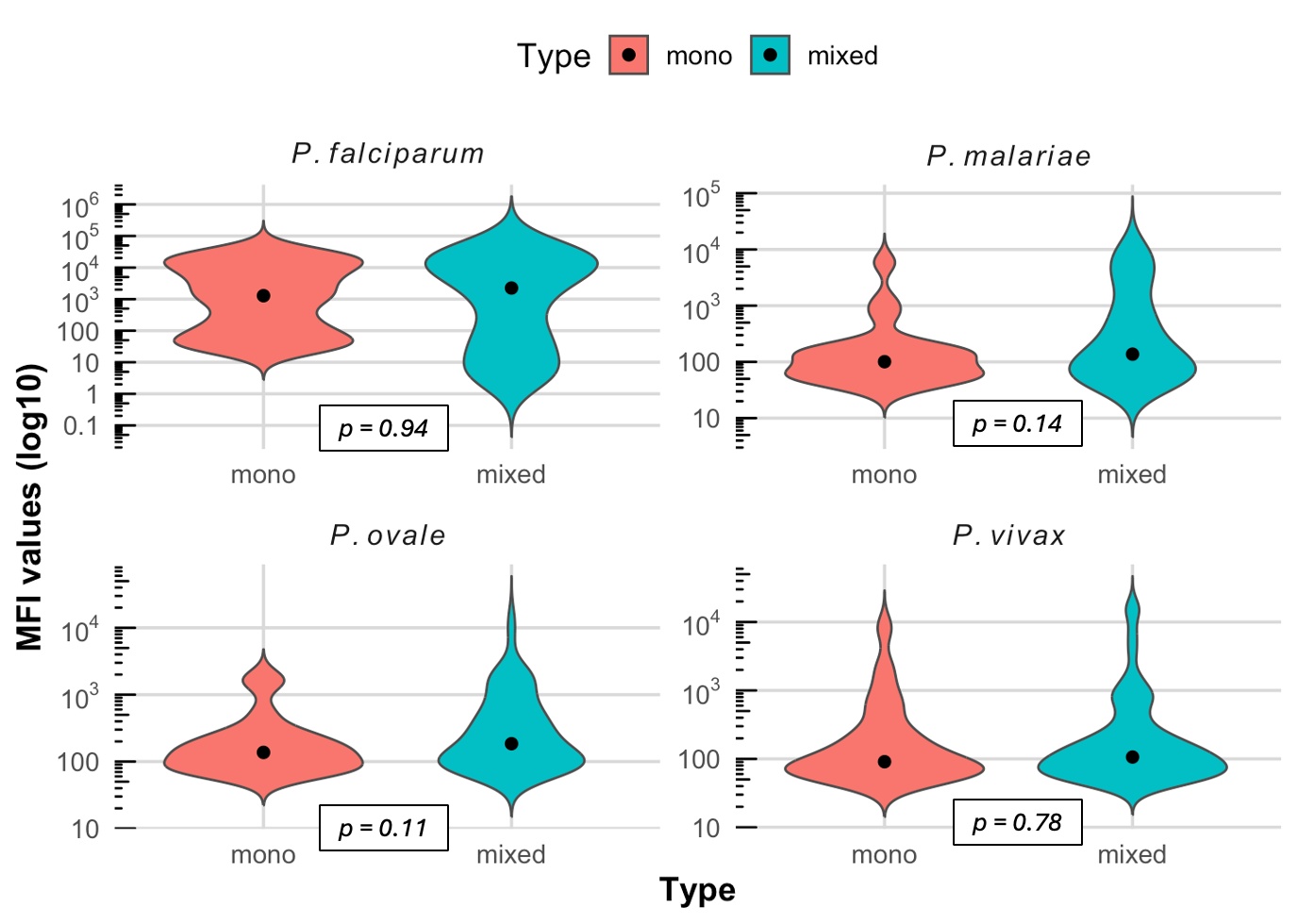

**Supplementary Figure 3. Distribution of MFI between individuals with exposure to 1 species (mono) compared to those with multiple exposures (mixed) in addition to the 1 species, using a 3SD cutoff.** Dots indicate median values for each sample. Non-parametric two-sided Mann–Whitney U (Wilcoxon rank‐sum) tests suggest no significant differences in distributions across all species (*p* > 0.05).

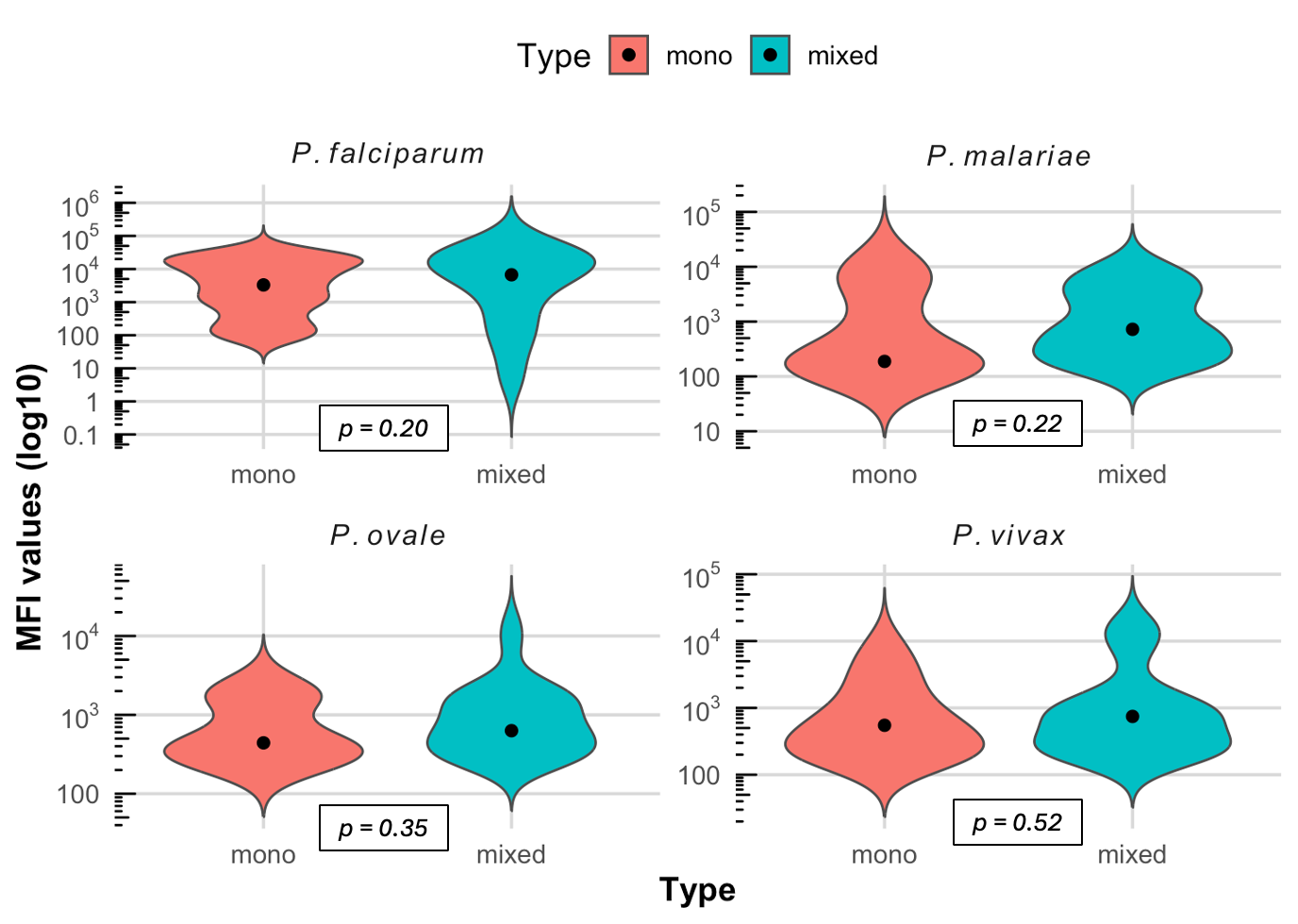

**Supplementary Figure 4. Distribution of MFI between individuals with exposure to 1 species (mono) compared to those with multiple exposures (mixed) using a 5SD cutoff.** Dots indicate median values for each sample. Non-parametric two-sided Mann–Whitney U (Wilcoxon rank‐sum) tests suggest no significant differences in distributions across all species (*p* > 0.05).

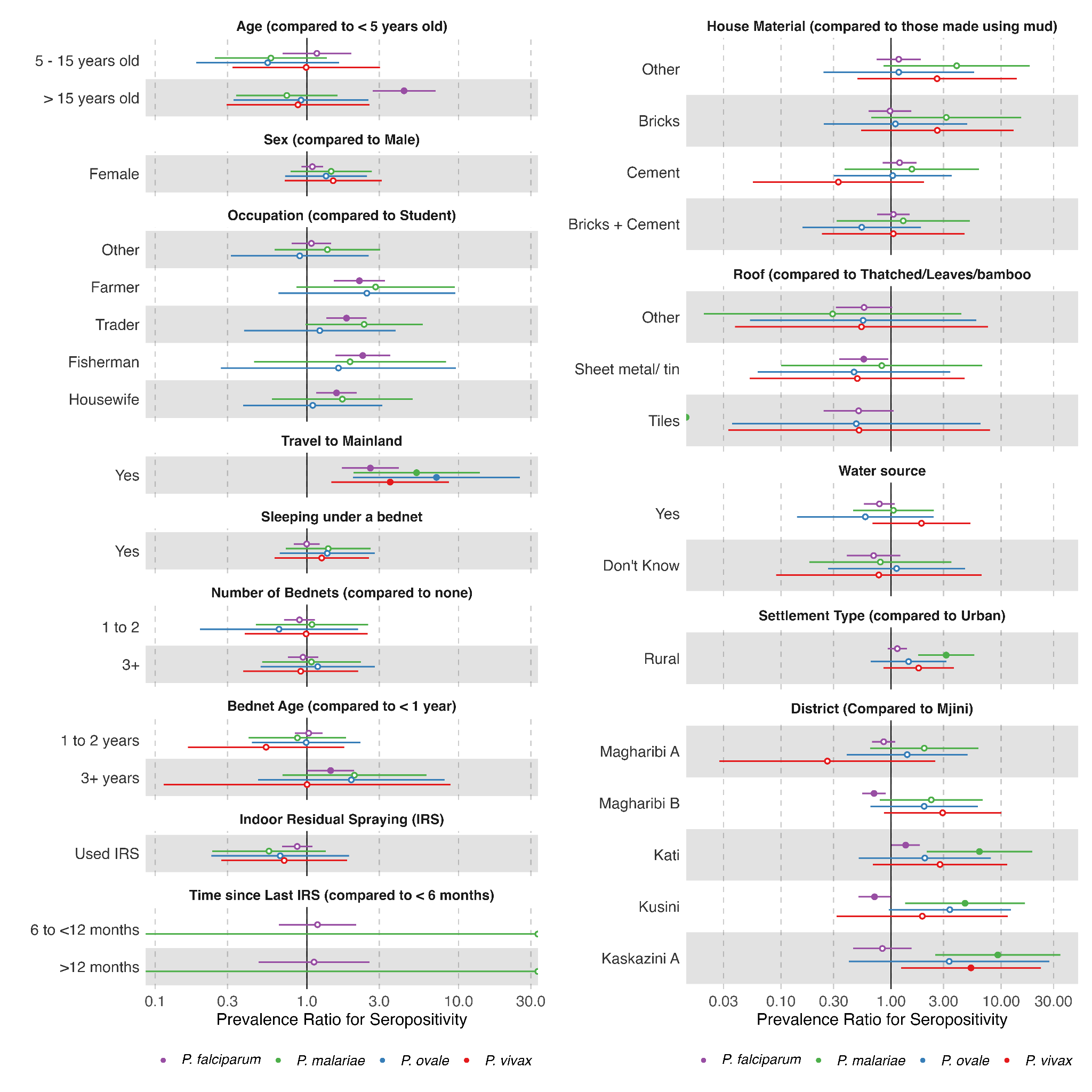

**Supplementary Figure 5. Factors associated with seropositivity for all four species using a 5SD cutoff.** Seroprevalence ratios were estimated by fitting robust poisson regression models using Generalized Estimating Equations (GEEs) to each malaria species, adjusting for individual age and clustering of individuals within the same households.
